# Tracking Immunity: Increased number of COVID-19 boosters increases the longevity of anti-RBD and anti-RBD neutralizing antibodies

**DOI:** 10.1101/2024.11.25.24317883

**Authors:** Ching-Wen Hou, Stacy Williams, Veronica Boyle, Alexa Roeder, Bradley Bobbett, Izamar Garcia, Giavanna Caruth, Mitch Magee, Yunro Chung, Douglas Lake, Joshua LaBaer, Vel Murugan

## Abstract

**Background/Objectives:** Since the World Health Organization declared COVID-19 a pandemic in March 2020, the virus has caused multiple waves of infection globally. Arizona State University (ASU), the largest four-year university in the United States, offers a uniquely diverse setting for assessing immunity within a large community. This study aimed to evaluate SARS-CoV-2 antibody seroprevalence and the effects of infection and vaccination three years into the pandemic.

**Methods:** A serosurvey was conducted at ASU from January 30 to February 3, 2023. Participants completed questionnaires about demographics, respiratory infection history, symptoms, and COVID-19 vaccination status. Blood samples were analyzed for anti-receptor binding domain (RBD) IgG and anti-nucleocapsid (NC) antibodies, offering a comprehensive view of immunity from both natural infection and vaccination.

**Results:** The seroprevalence of anti-RBD IgG antibodies was 96.2% (95% CI: 94.8%-97.2%), and 64.9% (95% CI: 61.9%-67.8%) of participants had anti-NC antibodies. Anti-RBD IgG levels correlated strongly with neutralizing antibody levels, and participants who received more vaccine doses showed higher levels of both anti-RBD IgG and neutralizing antibodies. Increasing number of exposures through vaccination and/or infection resulted in higher and long-lasting antibodies.

**Conclusion:** The high levels of anti-RBD antibodies observed reflect substantial vaccine uptake within this population. Ongoing vaccination efforts, especially as new variants emerge, are essential to maintaining protective antibody levels. These findings underscore the importance of sustained public health initiatives to support broad-based immunity and protection.

## 1. Introduction

Arizona State University (ASU) has the largest undergraduate population among all 4-year colleges in the United States [1]. ASU provides a unique and valuable setting to investigate the presence of COVID-19 antibodies within a large and diverse community of students, staff and faculty. ASU has notably diverse community [2], offering an opportunity to study how immunity has developed over time, particularly in the years following the WHO’s declaration of COVID-19 as a pandemic on March 11, 2020.

As of May 3, 2023, there have been 765 million reported cases of COVID-19 and 6.9 million reported deaths [2]. However, these figures likely underestimate the true infection rate due to factors like testing limitations and asymptomatic cases. Population-based serosurveys have become crucial in estimating the actual spread of SARS-CoV-2, revealing the extent of both past infections and vaccine-induced immunity. Studies have shown significant variation in seroprevalence rates depending on factors such as geographic location, population density, public health measures, and vaccination coverage [3]. For instance, a study assessed IgG antibody seroprevalence and risk factors for SARS-CoV-2 infection, showing an increase in seroprevalence from 28.5% to 71.5% between the first and second pandemic waves. Urban and rural areas with lower socioeconomic status had the highest seroprevalence (75.1%). These findings highlight the need for improved vaccination strategies to reach high-risk groups to enhance preparedness for future outbreaks[4]. Chen and colleagues found that the risk of infection was significantly higher among Black individuals (relative risk [RR] 2.70, 95% CI 2.30–3.18) and Asian individuals (RR 1.91, 95% CI 1.82–2.03) compared to White individuals. Additionally, the infection risk was elevated among working-age adults (20–64 years) in contrast to younger (<20 years) and older (≥65 years) age groups[5]. These studies provide valuable insights for guiding public health strategies and decision-making [6]. However, most of these studies were conducted during the first two years of the pandemic, from 2020 to 2022, leaving limited data from later seroprevalence studies while SARS-CoV-2 is still circulating.

To explore the prevalence of COVID-19 infections and vaccination coverage after multiple waves of the pandemic, we conducted a comprehensive serological survey in January 2023, roughly three years after the pandemic began. In our study, we employed various assays to measure antibodies targeting the RBD of the spike (S) and nucleocapsid (NC) proteins of the virus, along with neutralizing antibodies, which are key indicators of the immune system’s ability to fight off infection. By understanding the levels of these antibodies in the population, we can estimate for COVID-19 vaccination coverage, infection rate, and seroprevalence within the community.

## 2. Materials and Methods

### 2.1 Ethical Approval

The study was approved by ASU’s institutional Review Board (IRB) (STUDY00015522).

### 2.2 Study Design and Participants

Recruitment for this study was conducted through invitations, email announcements to the ASU community, and social media advertising, and potential participants were required to complete an electronic consent and a survey before giving biological specimens. The sample collection was extended for three days, from January 30 1 to February 3, 2023. Participants were initially invited via emails and social media channels. Individuals were eligible for inclusion if they were 18 years of age or older and were able to provide informed consent. 999 individuals, including ASU students and employees, who completed the screening, provided informed consent, and filled out the initial survey forms, were successfully recruited for the study. Participants were compensated for their time and efforts towards completing the survey and submitting samples. Individuals under 18, those unable to provide consent, pregnant women, or those weighing less than 110 lbs. at the time of the survey were excluded (Figure 1).

**Figure 1.**
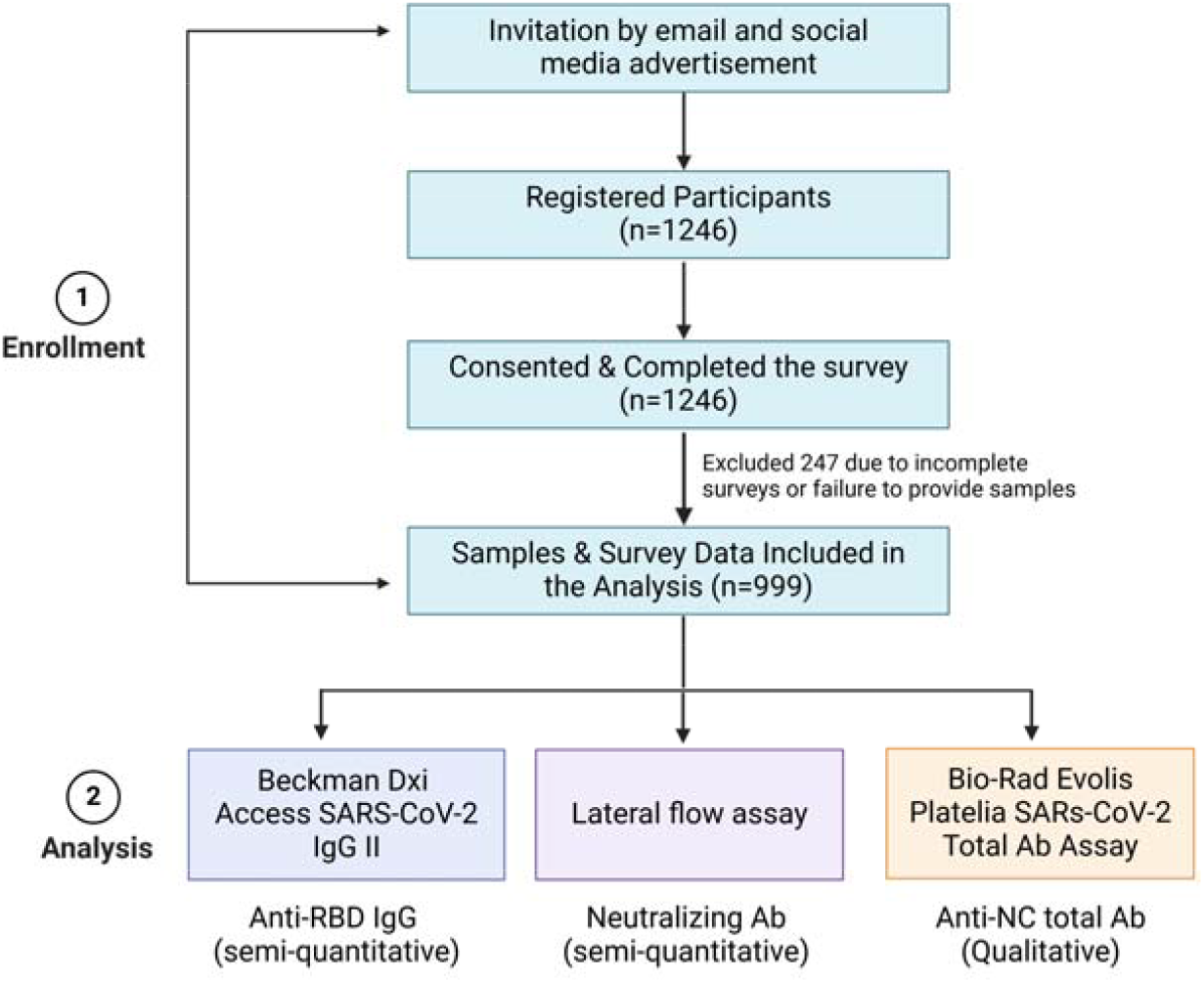
Flowchart of the study design. Recruitment, sample selection, and the assays/specimens used to detect antibody levels.

### 2.3 Survey Instruments

Demographic information, COVID-19 vaccination status, testing history, and symptoms were collected through a self-reported questionnaire. Participants voluntarily provided this information and were compensated after providing samples.

### 2.4 Blood Sample Collection

Blood samples were collected by trained phlebotomists at ASU using serum tubes (Cat #37988 from BD). Within 4 hours of collection, the samples were placed in a cooler for transportation to the laboratory. Upon arrival, the samples were centrifuged at 1300 g for 20 minutes to separate the serum. A total of 999 serum samples, along with their corresponding survey results, were included in the analysis.

### 2.5 Serology testing

In this survey, we used serological assays from two different platforms; 1) an EUA authorized, semi-quantitative Beckman Access SARS-CoV-2 IgG II assay to measure anti-RBD IgG antibodies; 2) an EUA authorized, qualitative Bio-Rad Platelia SARS-CoV-2 Total Ab ELISA assay to measure anti-NC antibodies.

The Access SARS-CoV-2 chemiluminescent IgG II, a semi-quantitative assay, used five different concentrations of calibrators and two concentrations of controls provided by the manufacturer to ensure reagent integrity and proper assay performance before sample analysis. The results were then compared to a cutoff value, expressed in arbitrary units (AU/mL), which was established during the instrument calibration process.

The interpretation of the Platelia SARS-CoV-2 Total Ab ELISA assay followed the manufacturer’s guidelines provided in the instructions for use (IFU). Values less than 0.8 were considered negative, values between 0.8 and 1.0 were categorized as equivocal, and values greater than 1.0 were considered positive.

### 2.6 Neutralization assay

The Lateral Flow Neutralizing Antibody (NAb) assay is designed to measure antibody levels that compete with ACE2 for binding to the receptor-binding domain (RBD) of SARS-CoV-2[7-9]. The test uses a single-port cassette containing a sample pad, nitrocellulose membrane, and conjugate pad with RBD-conjugated nanoshells. When neutralizing antibodies are present, they block RBD from binding to ACE2, resulting in a faint or absent test line. The test is semi-quantitative, and results can be read using either a scorecard or densitometer [8]. Univariate ROC analysis showed discrimination of neutralizing samples with a sensitivity and specificity of 0.9 and 1.0 respectively [8].

### 2.7 Statistical analysis

We performed descriptive statistics for demographic, vaccination-related, and self-reported previous infection variables. We then estimated the seroprevalences of anti-RBD IgG and anti-NC total antibodies and compared these by self-reported infection and vaccination statuses using two-sample proportional tests. Additionally, we estimated the seroprevalences of anti-RBD IgG and anti-NC total antibodies of participants who reported no previous infections by the number of vaccinations received. We used relative risk models to model the ratio of seroconversion probability as a linear function of the demographic variables. Due to the small sample size of those 65 years and older, we combined those individuals with participants 41 to 65 years of age. We subset the data by those who self-reported infection (n=529), with the seroconversion outcome variable being whether they were NC ELISA positive or negative. We also subset the data by those who were fully vaccinated (n=865), with the seroconversion outcome variable being whether they were positive or negative from the Beckman assay. We were also interested in determining if there was a relationship between the percentage of neutralizing antibodies against SARS-CoV-2 WT or Anti-RBD IgG levels and the number of vaccine doses. We further delved into the patterns exhibited by Anti-RBD IgG levels by looking at the duration between participants’ latest vaccination date and the collection date, dividing them into four categories: 0-6 months, 7-12 months, 13-18 months, and 19-24 months. Comparisons between vaccination doses and time frames were conducted using the Mann-Whitney test. We then assessed the percentage of neutralizing antibodies against SARS-CoV-2 WT to the Anti-RBD IgG levels using Spearman’s coefficient correlation. Finally, we performed linear regression to determine whether the rate at which Anti-RBD IgG levels and percentage of neutralizing antibodies decayed over the time following participants’ latest vaccination differed depending on the number of vaccines received. P-values less than 0.05 were considered statistically significant. R version 4.2.1 and GraphPad Prism 10.2.0 were used for statistical analysis.

## 3. Results

### 3.1. Demographic

A total of 999 participants were recruited for this serosurvey at Arizona State University, providing both saliva samples for qPCR diagnostic testing and blood donations. Among them, 585 (58.6%) were students, 376 (37.6%) were employees, 11 (1.1%) had an unspecified occupation, and 27 (2.7%) did not disclose their occupational status. Of the participants with occupation data, 526 (52.7%) were female, and 436 (43.6%) were male.

Regarding age distribution, 507 participants (50.8%) were between 18-25 years, 262 (26.2%) were aged 26-40 years, 190 (19.0%) were aged 41-65 years, 10 (1.0%) were over 65 years, and 30 (3.0%) did not report their age. Table 1 presents the demographic characteristics across these groups.

**Table 1.**
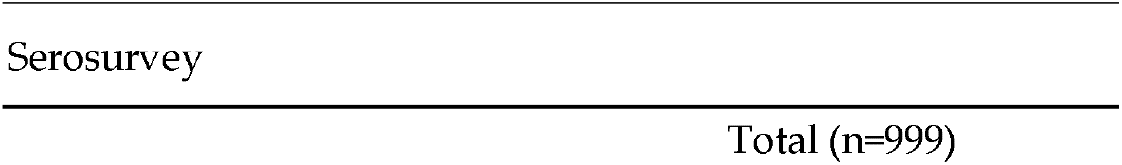
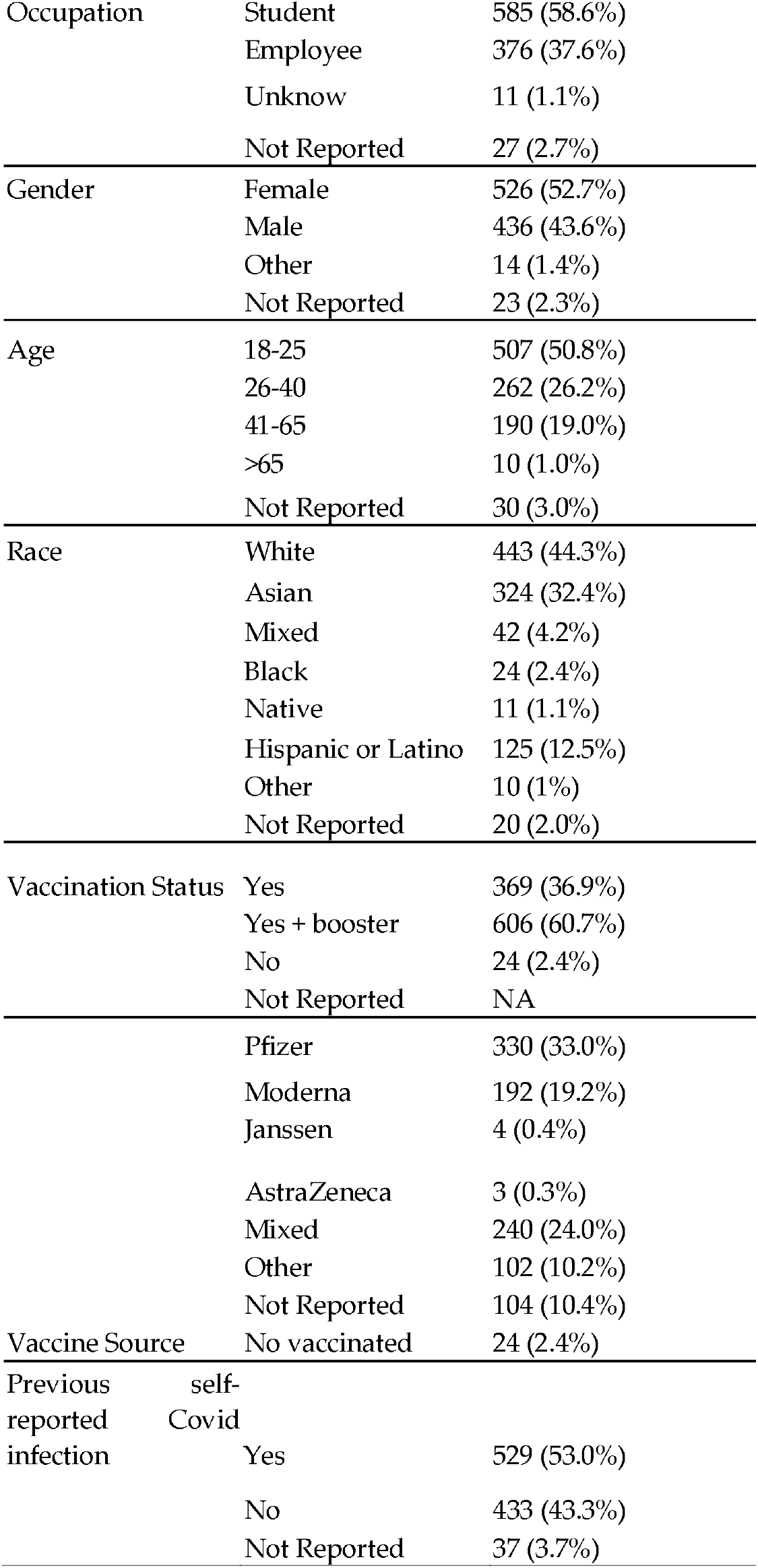
Demographic.

### 3.2 Self-reported COVID-19 infection and vaccine status

Out of the 999 participants, 53% (n=529) had previously tested positive for COVID-19, while 43.3% (n=433) had no record of a positive test (Table 1). Additionally, we examined the prevalence of PCR positivity among 999 asymptomatic students and employees from a university community on the sample collection day, finding a rate of 0.5% (n=5).

Regarding vaccination status, 999 participants provided information. Among these, 606 individuals (60.7%) reported being fully vaccinated with a booster dose, while 369 (36.9%) had received at least one dose of the vaccine but not a booster dose. A minority of 24 participants (2.4%) indicated they were unvaccinated. Concerning specific vaccines, the Pfizer vaccine was the most common, administered to 33% of participants (n=330/999), followed by Moderna, received by 19.2% (n=192/999). A smaller portion of the population received mixed vaccinations (24%), other vaccines (10.2%), remained unvaccinated (2.4%), or did not report their vaccination status (10.4%) (Table 1).

### 3.3 Seroprevalence

#### 3.3.1 SARS-CoV-2 RBD of spike IgG and NC total antibodies

The study analyzed 999 serum samples using the Access SARS-CoV-2 IgG II assay from Beckman Coulter to detect the presence of two antibody types: anti-RBD IgG antibodies and anti-nucleocapsid (anti-NC) total antibodies. The results revealed a high seroprevalence rate of anti-RBD antibodies at 96.2% (95% CI: 94.8% to 97.2%) and a lower rate for anti-NC antibodies at 64.9% (95% CI: 61.9% to 67.8%) (Table 2).

**Table 2.** Seroprevalence of anti-RBD and anti-NC antibodies.

| Antigen Detected | Ab Sub-Type | Manufacturer | Sample Type | Name of the test | Positives | Negative | Inconclusive |
| --- | --- | --- | --- | --- | --- | --- | --- |
| RBD | IgG | Beckman Coulter | Serum | Access SARS-CoV-2 IgG II (Semi-Quantitative) | 961 (96.2%)<br>95% CI: (94.8%, 97.2%) | 38 (3.8%)<br>95% CI: (2.8%, 5.2%) | 0 (0%)<br>95% CI: (0%, 0.4%) |
| Nucleocapsid | Total Ab | Bio-Rad | Serum | Platelia SARS-CoV-2 Total Ab Assay | 648 (64.9%)<br>95% CI: (61.9%, 67.8%) | 321 (32.1%)<br>95% CI: (29.3%, 35.1%) | 30 (3%)<br>95% CI: (2.1%, 4.3%) |

Of the surveyed participants, 206 reported no history of COVID-19 infection but had been vaccinated (excluding those who received AstraZeneca, Janssen, and mixed/other vaccines). Among these individuals, 198 (96.1%) tested positive for anti-RBD antibodies, while 74 (35.9%) were positive for anti-NC antibodies. In another group, among the 529 participants who reported a previous COVID-19 infection, irrespective of vaccination status, 404 (76.4%) tested positive for anti-NC antibodies (p<0.001, two-sample proportion test) (Table 3).

**Table 3.** Seroprevalence of anti-RBD and anti-NC antibodies in different cohorts.

| Cohort |  |  | RBD Protein IgG | Nucleocapsid Protein Total |
| --- | --- | --- | --- | --- |
| Infection | Vaccine | n | Beckman | ELISA |
| YES | YES | 514 | 507 (98.6%)<br>95% CI: (97.2%, 99.3%) | 394 (76.7%)<br>95% CI: (72.8%, 80.1%) |
|  | NO | 15 | 11 (73.3%)<br>95% CI: (48%, 89.1%) | 10 (66.7%)<br>95% CI: (41.7%, 84.8%) |
|  | NA | 0 | NA | NA |
| NO | YES | 206* | 198 (96.1%)<br>95% CI: (92.5%, 98%) | 74 (35.9%)<br>95% CI: (29.7%, 42.7%) |
|  | NO | 8 | 5 (62.5%)<br>95% CI: (30.6%, 86.3%) | 7 (87.5%)<br>95% CI: (52.9%, 97.8%) |
|  | NA | 0 | NA | NA |
| NA | YES | 36 | 35 (97.2%)<br>95% CI: (85.8%, 99.5%) | 26 (72.2%)<br>95% CI: (56%, 84.2%) |
|  | NO | 1 | 1 (100%)<br>95% CI: (20.7%, 100%) | 1 (100%)<br>95% CI: (20.7%, 100%) |

\*remove people who had received AstraZeneca, Janssen, and other/mixed vaccines

Regarding vaccination status, 10 participants who had no previous infection reported receiving a single vaccine dose, with an estimated anti-RBD seroprevalence of 60.0% (6/10; 95% CI: 31.3% to 83.2%). In contrast, the seroprevalence was notably higher at 88.9% (104/117; 95% CI: 81.9% to 93.4%) for those who received two doses. After receiving a booster dose, seroprevalence of anti-RBD antibodies remained consistently high at 96.4% (135/140; 95% CI: 91.9% to 98.5%). Interestingly, seroprevalence of anti-NC antibodies decreased with an increase in vaccine doses (Table 4). Additionally, among those vaccinated without prior infection who tested negative for anti-NC antibodies, the anti-RBD seroprevalence reached 100% following five vaccine doses, with a significant rise observed from three to four doses (p<0.001) (Table 4).

**Table 4.**
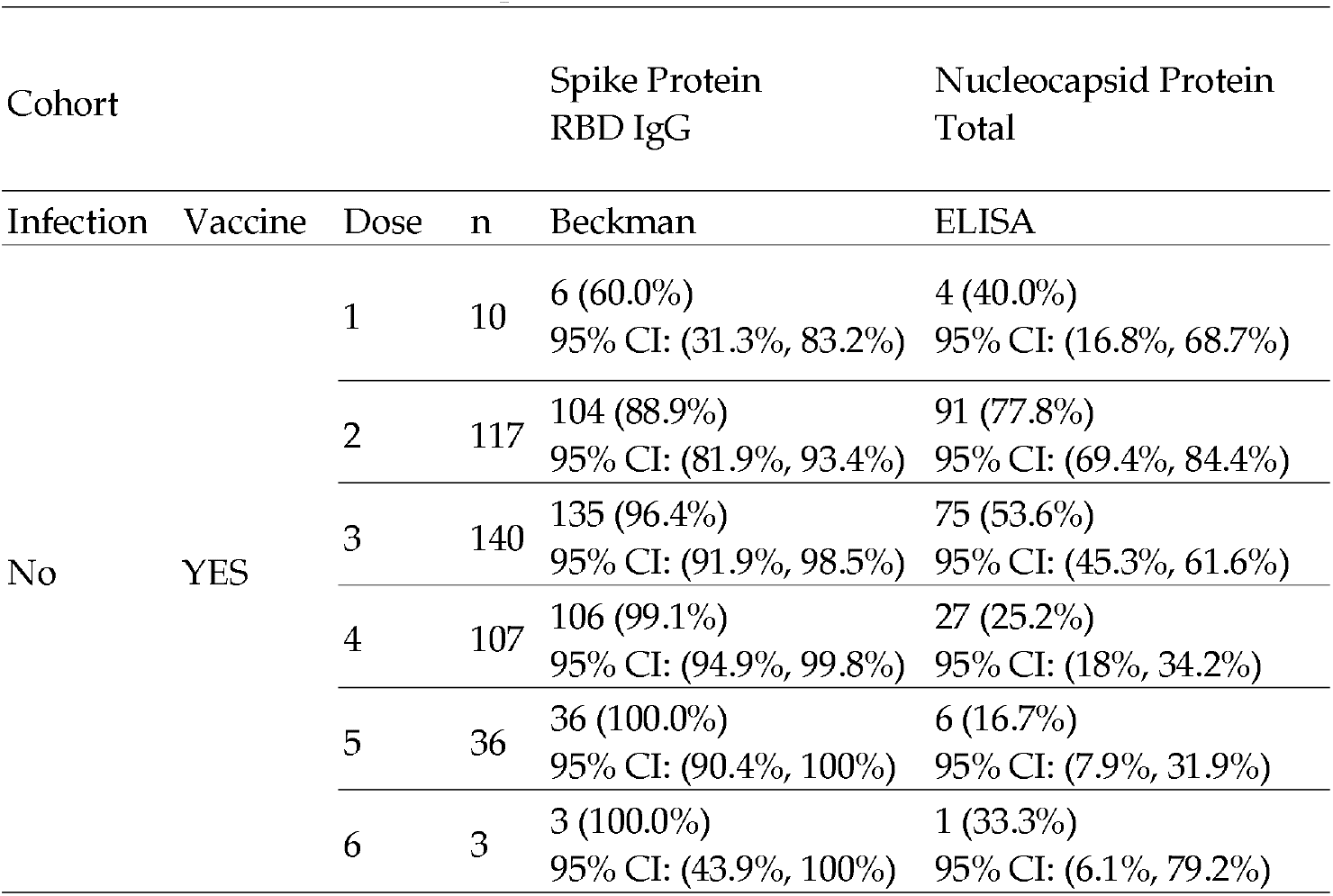
Seroprevalence of anti-RBD and anti-NC antibodies compared to the number of doses.

When comparing anti-RBD IgG seroprevalence by vaccine type, the highest seroprevalence was among recipients of the Moderna vaccine (97.5%, 79/81; 95% CI: 91.4% to 99.3%), followed by Pfizer (95.2%, 119/125; 95% CI: 89.9% to 97.8%), and those with mixed/other vaccines (93.5%, 158/169; 95% CI: 88.7% to 96.3%) (Table 5).

**Table 5.** Anti-RBD seroprevalence in participants received vaccine without previous infection based on the self-reported data.

| Cohort |  |  |  | Spike Protein<br>RBD IgG |
| --- | --- | --- | --- | --- |
| Infection | Vaccine | Source | n | Beckman |
| No | YES | Pfizer | 125 | 119 (95.2%)<br>95% CI: (89.9%, 97.8%) |
|  |  | Moderna | 81 | 79 (97.5%)<br>95% CI: (91.4%, 99.3%) |
|  |  | Mixed/Others* | 169 | 158 (93.5%)<br>95% CI: (88.7%, 96.3%) |

\*received AstraZeneca, Janssen, and other/mixed vaccines

### 3.4 Seroconversion and demographic variables

In examining seroconversion rates, we assessed demographic factors—including race, gender, age, employment status, and vaccine type—among the ASU community. Seroconversion indicates the appearance of detectable antibodies in the blood post-infection or vaccination. Our analysis found no significant differences in the production of anti-RBD or anti-NC antibodies across different races, age groups, genders, or employment statuses following vaccination or self-reported exposure to SARS-CoV-2 (Table S1).

### 3.5 Anti-RBD IgG antibody levels after vaccination

Previous studies have reported a decrease in anti-SARS-CoV-2 antibody levels within the first six months following COVID vaccination[10-12]. In our study, among participants who received the vaccine without a prior infection based on self-reported data and tested negative for anti-NC total antibodies, we similarly observed a scientific decrease in anti-RBD antibody and neutralizing antibody levels six months post-vaccination (Figure 2A-B). Our findings revealed that antibodies remained detectable even 24 months after vaccination. A significant decline was noted between the 0– 6-month and 7–12-month post-vaccination periods in this serosurvey (p=0.0002), with levels dropping from 128.2 AU/mL to 57.44 AU/ mL. Antibody levels continued to decline, reaching 36.22 AU/mL for participants 13–18 months post-vaccination.

**Figure 2.**
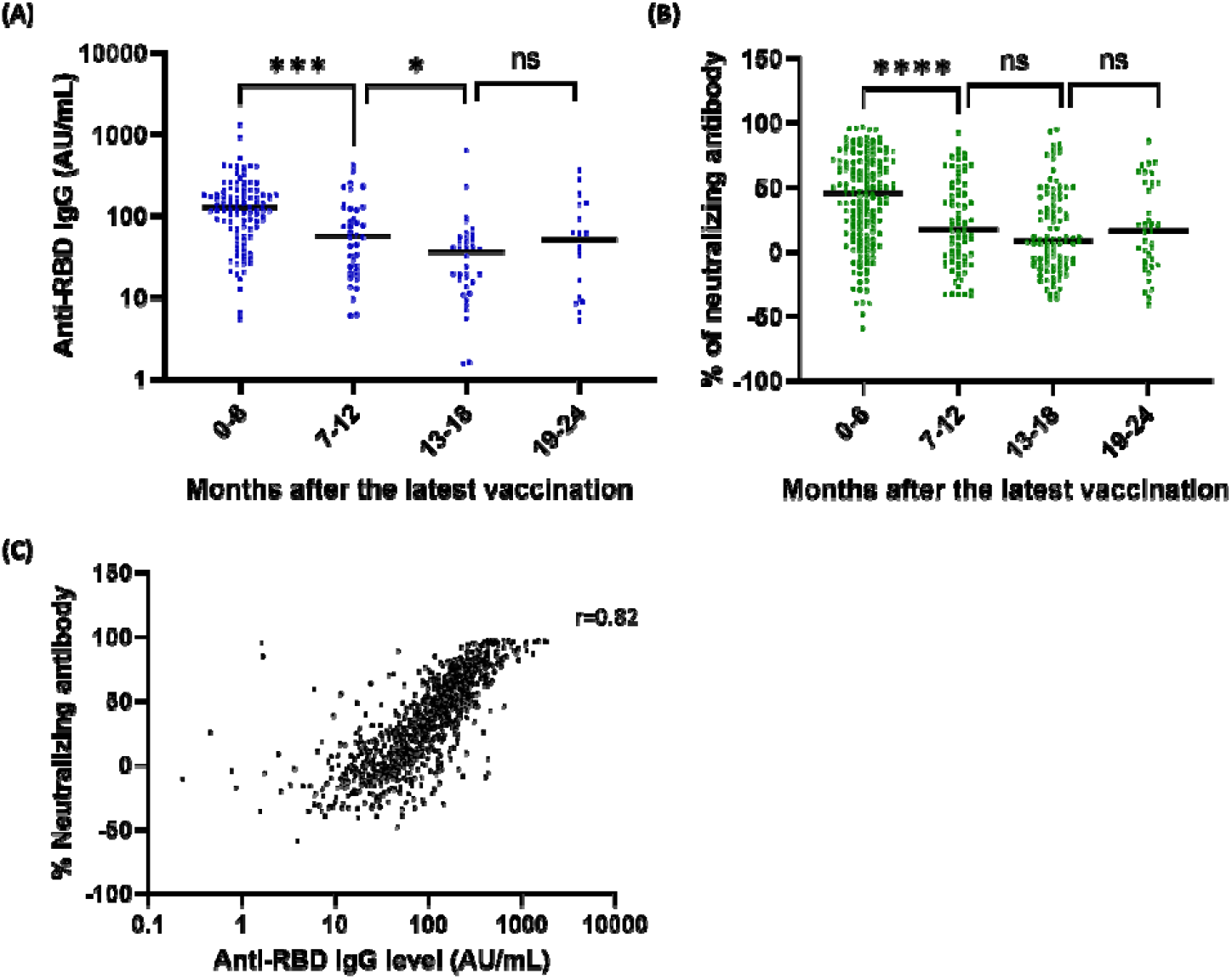
Antibody response in participants after the latest vaccination and the correlation between the levels of anti-RBD IgG and neutralizing antibodies. (A) Anti-RBD antibodies were measured using Beckman immunoassay and (B) Neutralizing antibody was measured using lateral flow assay in participants who had previous COVID-19 vaccines without self-reported prior infection. (C) Correlation between levels of anti-SARS-CoV-2 RBD IgG and neutralizing against WT.

We also observed that anti-RBD IgG levels and neutralizing antibody increased with the number of vaccine doses (Figure 3A & 3B) and decayed more slowly among participants who received ≥ 4 doses compared to those who received ≤ 3 doses (Figure 3C & 3D).

**Figure 3.**
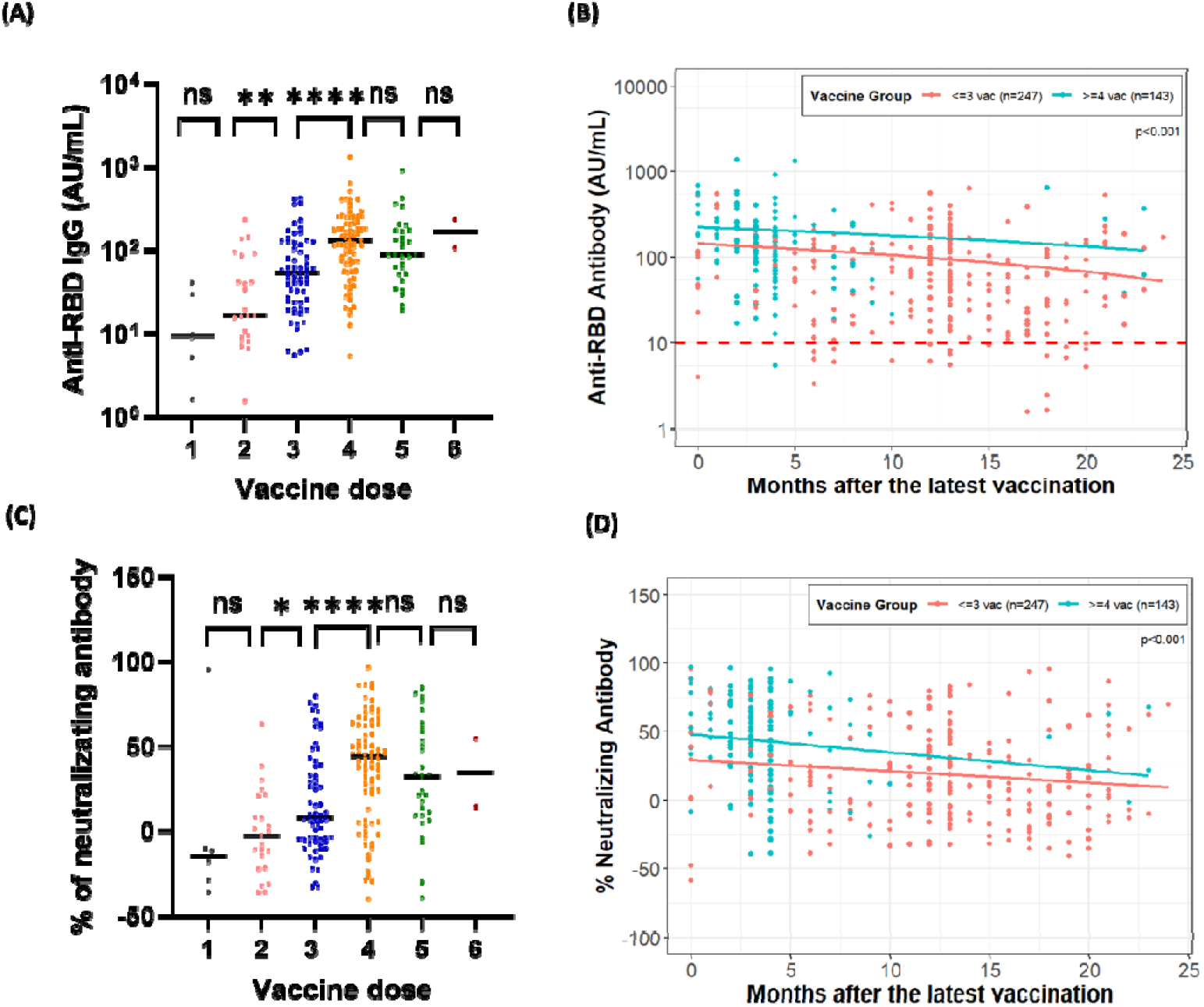
Anti-RBD antibody and percent neutralizing antibody response in participants who had rec vaccination without previous infection and tested negative for anti-NC total antibodies after vaccine dose. (A) RBD antibody was measured by the Beckman assay (B) Anti-RBD antibody decay (C) Percent of neutralizing antibody was measured using a lateral flow assay (D) Neutralizing antibody decay.

Next, if we consider an infection as an additional SARS-CoV-2 antigen exposure alongside vaccination, we calculated the overall impact of SARs-CoV-2 antigen exposure (from infection or vaccination) on the decay of anti-RBD antibodies. Figure S1 showed that participants with four or more exposures to the COVID-19 antigen (from infection or vaccination) exhibited significantly slower decay rates of RBD antibodies and neutralizing antibodies (p < 0.001) compared to those with fewer than three exposures. The results showed that greater antigen exposure correlates with a slower rate of antibody decay.

### 3.6 Comparison of Anti-RBD IgG and neutralizing antibody responses

Finally, we compared responses between anti-RBD IgG and neutralizing antibodies. Using a rapid lateral flow assay [8], we observed that neutralizing antibodies against the SARS-CoV-2 wild-type strain rose consistently with additional vaccine doses, with a notable increase from three to four doses (p<0.0001) (Figure 3A). The correlation between neutralizing antibody and anti-RBD IgG levels was strong, with a Spearman’s correlation coefficient of 0.82 (95% CI: 0.80–0.84; p<0.0001) (Figure 2C).

## 4. Discussion

Conducting a serosurvey three years into the COVID-19 pandemic is essential for understanding the longer-term dynamics of immunity within the population. With the virus evolving and new variants emerging, immunity— whether from natural infection or vaccination—has shifted over time. A serosurvey provides critical insights into antibody prevalence, allowing us to evaluate the durability of vaccine effectiveness, the scope of natural immunity, and the potential need for booster doses or updated vaccines. By assessing the current status of population immunity, we can better prepare for future outbreaks and manage the virus as it potentially transitions to an endemic phase.

This study observed a seroprevalence rate of 96.2% for anti-RBD IgG antibodies and 64.9% for anti-NC total antibodies in the local community after three years of the pandemic (Table 2). The seroprevalence rate of anti-NC total antibodies continued to increase compared to studies conducted in September 2021 [13] and March 2022 [14] at ASU. The seroprevalence of anti-RBD IgG antibodies observed in this study was higher than that reported in the September 2021 [13] study and was comparable to the findings from March 2022 [14]. The observed changes in seroprevalence rates of anti-RBD and NC antibodies suggest a few key points.

The sustained seroprevalence of anti-RBD IgG antibodies observed in our study, which remained high compared to the September 2021 study and was consistent with the March 2022 findings, suggests that the immunity provided by vaccination has been maintained over time. This ongoing immunity could be attributed to booster doses, as 60.7% of participants in this study reported having received at least one booster (Table 1), contributing to stable levels of anti-RBD antibodies within the population. Similarly, a sero-monitoring study in New York City, analyzing data from over 55,000 individuals across five pandemic waves, found a steady rise in antibody levels, especially after vaccine boosters and breakthrough infection, with seroprevalence surpassing 90% by July 2022. These findings demonstrate the lasting impact of vaccination and booster doses in maintaining immunity as the pandemic shifted to an endemic stage [15]. However, not all populations worldwide exhibit high seroprevalence after multiple COVID-19 waves, as highlighted in this study. An analysis of 247 studies involving 757,075 children from 70 countries revealed that seroprevalence increased over time, from 7.3% (5.8–9.1%) during the first wave to 37.6% (18.1–59.4%) in the fifth wave and 56.6% (52.8–60.5%) in the sixth wave. The highest rates were observed in South-East Asia (17.9–81.8%) and Africa (17.2– 66.1%), while the Western Pacific region reported the lowest rates (0.01–1.01%). [16]. Similarly, a study in South Africa reported seroprevalence levels of 60% in a rural community and 70% in an urban community following the third wave of SARS-CoV-2 infections [17]. These findings emphasize the urgent need to enhance vaccine access and coverage, particularly in developing countries and among minority ethnic groups.

Our results also showed no significant differences in the production of anti-RBD and anti-NC antibodies across different races, age groups, genders, or employment statuses, which contrasts with findings in other studies [18,19]. Those studies observed age- or gender-dependent antibody responses in healthcare workers following mRNA SARS-CoV-2 vaccination. The discrepancy may stem from differences in study design: their studies were longitudinal and focused on healthcare workers, whereas ours is a cross-sectional study primarily involving students. When considering all variables, individuals who received mRNA vaccines did not exhibit higher seroconversion rates compared to those who received other vaccine types (Supplementary Table 1). However, when considering only vaccine type, individuals who received mRNA vaccines exhibited higher seroconversion rates, consistent with other findings [20,21] (Table 5).

In this study, we observed a significant decline in anti-RBD and neutralizing anti-body levels during the first six months post-vaccination (Figure 2A-B). Notably, anti-RBD IgG levels and neutralizing antibody titers increased with the number of vaccine doses administered (Figure 3A and 3C). Moreover, participants who received ≥4 doses exhibited significantly slower decay of both anti-RBD and neutralizing antibodies compared to those who received ≤3 doses (Figure 3B and 3D). These findings aligned with existing literature [22], which showed that antibody responses to SARS-CoV-2 mRNA vaccination undergo a rapid waning phase initially, followed by stabilization around 7 to 9 months post-vaccination. Furthermore, booster vaccinations eliminated differences in antibody levels between individuals with and without hybrid immunity. Breakthrough infections in previously naïve individuals acted as effective boosters, raising antibody levels to titers comparable to those observed after an additional vaccine dose.

We also demonstrated a strong correlation between anti-RBD IgG antibody levels and neutralizing antibody levels against SARS-CoV-2 WT, which is consistent with findings from other studies [23-25]. However, our study did not detect neutralizing antibodies against other variants, so we could not assess the correlation between them and anti-RBD IgG antibodies. The correlation might change with different mutations, especially after multiple waves. On the other hand, the results may not change significantly due to the phenomenon known as “original antigenic sin” in which an immunized host continues to produce antibodies against the first immunogen even after infection or immunization with different variants [26,27].

Finally, the continued increase in the seroprevalence of anti-NC total antibodies observed in this study compared to the first two years’ surveys[13,14] indicates ongoing exposure to the SARS-CoV-2 virus. Since anti-NC antibodies typically arise from natural infection rather than vaccination, this suggests that a significant portion of the population has been infected with SARS-CoV-2, leading to a natural increase in these antibodies over time. Notably, 35.9% of participants who reported no previous COVID-19 infection tested positive for anti-nucleocapsid antibodies in this study (Table 3). This suggests that a significant number of asymptomatic or mildly affected COVID-19 cases may go undetected, potentially impacting public health strategies.

Additionally, we also observed that the seroprevalence of anti-RBD IgG antibodies increased while the seroprevalence of anti-NC antibodies decreased with the number of vaccine doses received (Table 4). Although this could suggest that participants who received updated COVID-19 vaccines could be protected from infection, SARS-CoV-2 continues to spread as it explores its evolutionary space.

## 5. Conclusions

In conclusion, the high prevalence of anti-RBD antibodies in the ASU community as of January 2023 suggests a level of immunity closer to herd immunity may have been achieved by the third year of the pandemic, but it is unlikely vaccines can keep pace with the evolution of the virus. Therefore, these results underscore the critical role of vaccination and booster doses in sustaining immunity to protect against severe disease and hospitalization, and highlight the importance of ongoing public health measures to maintain population protection against future outbreaks.

## Data Availability

All data produced in the present study are available upon reasonable request to the authors.

## Supplementary Materials

The following supporting information can be downloaded at: www.mdpi.com/xxx/s1, Table S1: Seroconversion by race, age, gender, employment status, and the types of vaccines; Figure S1: Anti-RBD antibody levels and neutralizing antibody percentages in participants exposed to SARS-CoV-2 antigens through infection or vaccination.

## Author Contributions

Conceptualization, V.M., D.L., M.M. and J.L.; methodology, V.M., D.L., J.L.,C.H., B.B., I.G., G.C., V.B. and A.R.; software, C.H., A.R. and S.W.; formal analysis, S.W. and Y.C.; investigation, V.M. and J.L.; writing—original draft preparation, C.H.; writing—review and editing, S.W., Y.C., A.R., D.L., J.L and V.M.; visualization, V.M.; supervision, V.M., J.L., and D.L. All authors have read and agreed to the published version of the manuscript.

## Institutional Review Board Statement

The study was conducted in accordance with the Declaration of Helsinki and approved by the Institutional Review Board of Arizona State University (protocol code STUDY00015522 and date of approval).” for studies involving humans.

## Informed Consent Statement

Informed consent was obtained from all subjects involved in the study. Written informed consent has been obtained from the patient(s) to publish this paper.

## Data Availability Statement

The dataset for this study can be found at DOI: 10.5061/dryad.1vhhmgr49..

## Acknowledgments

We would like to thank ASU EHS team for their logistical support and IBC oversight, communication team for their support in communicating study materials to ASU community, members of our clinical coordination team Lucy Sublasky-Rodriguez, Kelly Conard, Kylee Taylor and Eleni Katergaris for their help in sample accessioning, and Scarlett Goins-Heisler and Harrison Bell for their help in processing the blood tubes during the day of the serosurvey. We thank Steven Winn, and Jessica Lukosus for their help in managing the event and procurement of supplies. We also thank Funding provided by Arizona State University Knowledge Enterprise.

