## Supplemental Figure 1 and Table 1 for "Tracking Immunity: Increased number of COVID-19 boosters increases the longevity of anti-RBD and anti-RBD neutralizing antibodies"

**Table S1:** Seroconversion by race, age, gender, employment status, and the types of vaccines

| **Variable** | **Comparison** | **Anti-RBD Antibody**  **(Access SARS-CoV-2 IgG II)** | | | | **Anti-NC antibody**  **(Platelia NC total Ab)** | | | |
| --- | --- | --- | --- | --- | --- | --- | --- | --- | --- |
|  |  | **n** | **PR** | **95% CI** | **P-value** | **n** | **PR** | **95% CI** | **P-value** |
| Race | White vs Other | 368 vs 172 | 1.00 | (0.93, 1.08) | 0.93 | 229 vs 114 | 0.95 | (0.83, 1.09) | 0.50 |
|  | Asian vs Other | 268 vs 172 | 0.96 | (0.84, 1.09) | 0.53 | 134 vs 114 | 0.94 | (0.81, 1.08) | 0.35 |
|  | White vs Asian | 368 vs 268 | 1.05 | (0.92, 1.19) | 0.50 | 229 vs 134 | 1.02 | (0.90, 1.15) | 0.74 |
| Age | 26-40 vs 18-25 | 224 vs 373 | 1.02 | (0.91, 1.14) | 0.72 | 130 vs 191 | 0.91 | (0.79, 1.04) | 0.16 |
|  | 41+ vs 18-25 | 161 vs 373 | 1.00 | (0.85, 1.18) | 0.99 | 84 vs 191 | 1.07 | (0.89, 1.29) | 0.47 |
|  | 41+ vs 26-40 | 161 vs 224 | 0.98 | (0.86, 1.12) | 0.75 | 84 vs 130 | 1.18 | (0.99, 1.41) | 0.06 |
| Gender | Male vs Female | 366 vs 442 | 0.99 | (0.94, 1.05) | 0.84 | 211 vs 266 | 0.97 | (0.87, 1.07) | 0.52 |
| Employment Status | Student vs Employee | 487 vs 321 | 0.99 | (0.85, 1.14) | 0.85 | 278 vs 199 | 1.12 | (0.95, 1.31) | 0.17 |
| Vaccine Group^#^ | mRNA vaccine vs Other vaccine | 482 vs 326 | 0.99 | (0.88, 1.12) | 0.92 | 270 vs 157 | 0.91 | (0.82, 1.01) | 0.07 |
|  | Unvaccinated vs Other vaccine | N/A | N/A | N/A | N/A | 50 vs 157 | 1.06 | (0.72, 1.55) | 0.78 |
|  | mRNA vaccine vs Unvaccinated | N/A | N/A | N/A | N/A | 270 vs 50 | 0.86 | (0.58, 1.26) | 0.44 |

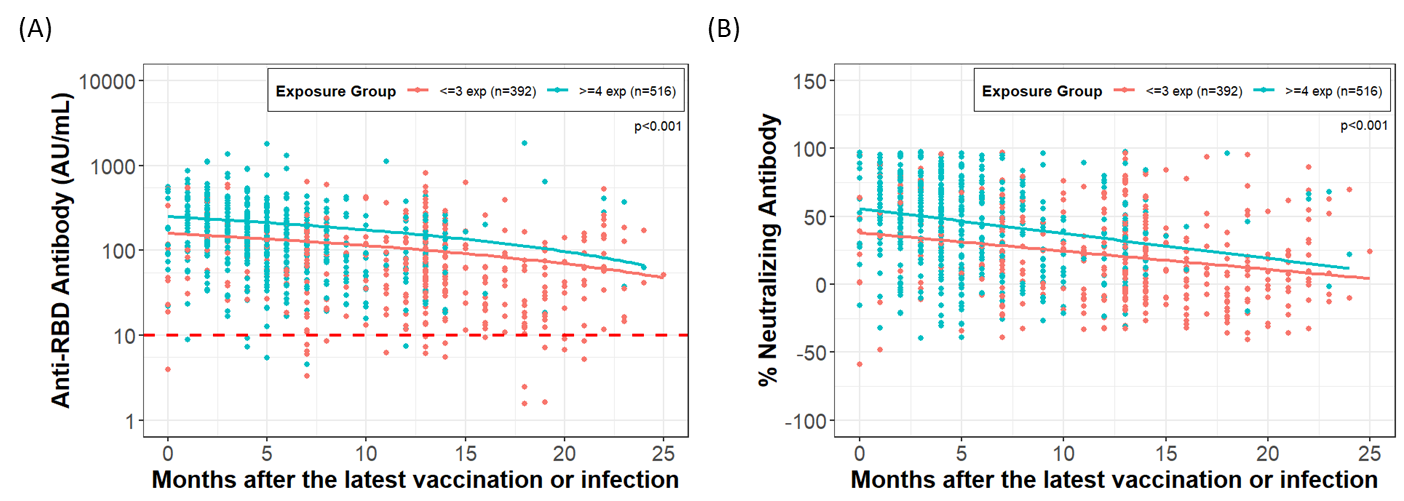

**Figure S1.** Anti-RBD antibody levels and neutralizing antibody percentages in participants exposed to SARS-CoV-2 antigens through infection or vaccination. (A) Anti-RBD antibody decay (B) Neutralizing antibody decay.
